# Increase in ventilatory ratio indicates progressive alveolar damage and suggests poor prognosis in severe COVID-19: A single-center retrospective observational study

**DOI:** 10.1101/2021.07.20.21260754

**Authors:** Natsuko Kaku, Yu Nakagama, Michinori Shirano, Sari Shinomiya, Kazuhiro Shimazu, Katsuaki Yamazaki, Yoshito Maehata, Ryo Morita, Yuko Nitahara, Hiromasa Yamamoto, Yasumitsu Mizobata, Yasutoshi Kido

## Abstract

**Background:** The symptoms of severe COVID-19 are complex and wide-ranging even in intensive care unit (ICU) patients, who may successfully discontinue respiratory support in a short period or conversely require prolonged respiratory support. Damage in the lungs of COVID-19 patients is characterized pathologically as diffuse alveolar damage, the degree of which correlates with the severity of the disease. We hypothesized that the ventilatory ratio (VR), a surrogate parameter for the dead space fraction, might stratify the severity of COVID-19 and predict the successful discontinuation of respiratory support.

**Methods:** Forty COVID-19 patients in our ICU were enrolled in this study. Respiratory variables were collected from 2 hours (day 0) after the initiation of respiratory support. We monitored the longitudinal values of VR and other respiratory parameters for 28 days. Patients successfully discontinued from respiratory support by day 28 of ICU stay were defined as the successfully discontinued group, while those who died or failed to discontinue were defined as the failed to discontinue group. VR and other respiratory parameters were compared between these groups.

**Results:** Except for advanced age, prolonged ventilation period, and higher mortality in the failed to discontinue group, there were no significant differences between the groups in terms of any other background or respiratory parameter at 2 hours (day 0) after initiation of respiratory support. Longitudinal VR monitoring revealed significantly higher VR values in the failed to discontinue group than the successfully discontinued group on day 4 of respiratory support. Upon predicting the failure to discontinue respiratory support, the area under the receiver operating characteristic curve of VR values on day 4 of respiratory support was 0.748. A threshold of 1.56 achieved the highest predictive performance with a sensitivity of 0.667 and a specificity of 0.762. This threshold enabled the prediction of the successfully discontinued outcome at 0.810 of the negative predictive value.

**Conclusions:** Elevated VR values on day 4 of respiratory support were predictive of successful discontinuation of respiratory support in patients with severe COVID-19. Longitudinal VR values after initiation of respiratory support can be used as a practical index to stratify severe COVID-19.

## Background

COVID-19, an infectious disease caused by severe acute respiratory syndrome coronavirus 2 (SARS-CoV-2), that emerged at the end of 2019, has a broad spectrum of manifestations from asymptomatic to critical illness. Even in the case of critically ill patients who are admitted to intensive care units (ICUs), some improve after only a few days of respiratory support, whereas others require prolonged respiratory support resulting in poor prognosis.

Similar to other lethal human coronaviruses such as SARS-CoV and MERS-CoV, the SARS-CoV-2 virus enters host cells, harnessing its spike protein to angiotensin-converting enzyme 2 (ACE2) receptor on the surface of the host cell (1). ACE2 is highly expressed in lung alveolar cells and bronchial epithelium, where the virus enters initially and proliferates (2). Therefore, the principal pathological manifestation of COVID-19 is considered to be alveolar damage, and the severity may depend on the progress of diffuse alveolar damage (DAD) (3,4) which is a typical feature of acute respiratory distress syndrome (ARDS) (5). However, the lack of appropriate indices for real-time monitoring of DAD is currently limiting stratification of patients who are critically ill with COVID-19.

DAD reduces effective gas exchange between alveolar epithelial cells and the pulmonary capillaries, thus increasing the functional dead space. Dead space fraction (V_D_/V_T_) is the most crucial factor dictating respiratory adequacy. Several studies have demonstrated the importance of V_D_/V_T_ in ARDS patients, in terms of both prognostication and disease progression (6). Despite its effectiveness, V_D_/V_T_ is seldom measured in usual ICU practice because of the complexity of calculation and the associated equipment costs (6).

Ventilatory ratio (VR) is a simple bedside index that has recently been proposed as a surrogate parameter for V_D_/V_T_ (6,7). It is easily calculated using minute ventilation and arterial partial pressure of carbon dioxide (PaCO_2_), as follows (8): 

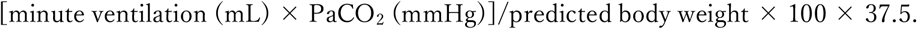

Sinha et al. have reported high VR as an independent predictor of mortality in two distinct ARDS randomized controlled trials (9,10), and have confirmed a significant correlation between VR and V_D_/V_T_ in ARDS (8).

Therefore, we monitored longitudinal VR in patients with severe COVID-19 who required respiratory support in our ICU with the aim of determining whether VR could stratify the severity of COVID-19 and predicting successfully discontinued prognosis.

## Methods

### Study design

This is a retrospective, single-center, observational study that enrolled patients with severe COVID-19 admitted into our ICU from February 29 to November 20, 2020. To identify the relationship between patient characteristics and prolonged respiratory support, the patients were stratified into two groups based on respiratory support dependency for the observed 28 days because the failure to discontinue respiratory support during the initial 28 days is known to have a strong correlation with mortality (11). In the observation period, patients who could not discontinue respiratory support were included to the failed to discontinue group and patients who could successfully discontinue respiratory support were included to the successfully discontinued group.

### Data collection

Data from patients’ electronic medical records were collected and reviewed by physicians trained in critical care. The inclusion criteria were age ≧18 years; SARS-CoV-2 infection confirmed by PCR or antigen test; and ICU admission for respiratory support, defined as the use of invasive mechanical ventilation. Of 45 eligible patients, 5 who were not ventilated at our hospital were excluded, and a total of 40 patients were included in the study.

The recorded data included patients demographics (age, gender, body mass index (BMI), comorbidities), disease course (days from onset to O_2_ requirement, days from onset to respiratory support, days of respiratory support, ICU mortality), laboratory and ventilatory parameters (minute ventilation (MV), tidal volume (TV), set PEEP, plateau pressure (Pplat), static lung compliance (Cstat), PaCO_2_, PaCO_2_/FiO_2_ (P/F) ratio, and VR). A complete data set was obtained 2 hours (day 0) after the patients was placed on respiratory support. VR and Cstat were calculated as follows:

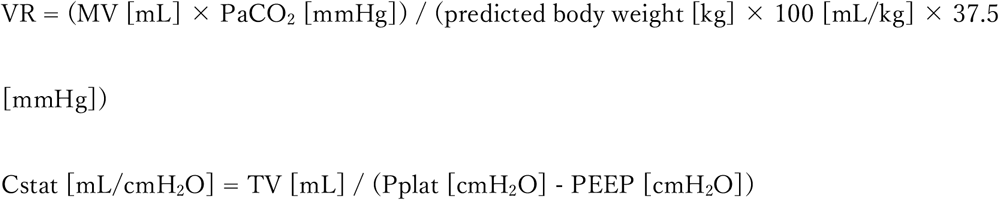

### Statistical analysis

All analyses were performed with GraphPad Prism 9.1.1 (GraphPad Software, Inc., San Diego, CA). Variables were compared between the groups using Mann-Whitney test for continuous variables and Chi-square test for binary data. To assess differences in longitudinal VR and its factors between the groups, VR values after starting respiratory support were plotted and analyzed by Mann-Whitney test. To validate whether the increased VR can become a prognostic index for successfully discontinued from respiratory support in severe COVID-19, we performed receiver operating characteristic (ROC) curves analysis of VR values on days 0-4 after respiratory support. All time points to events were defined from 2 hours (day 0) after initiation of invasive respiratory support. Missing data were not imputed. Days of respiratory support of patients who prolonged respiratory support longer than the observation period were deemed 28. Descriptive variables were expressed as the percentage or median and interquartile range (IQR), as appropriate. All tests were two-sided, and a *p*-value < 0.05 was considered statistically significant.

## Results

Among 45 patients admitted to our ICU between 29 Feb and 20 Nov 2020, 40 patients who met the inclusion criteria were enrolled in this study and followed for at least 28 days. The patient demographics, disease course, laboratory and respiratory parameters are listed in Table 1. Time from symptom onset to initiation of oxygen therapy was 7 days (IQR: 6–9 days) and that to initiation of respiratory support was 8 days (IQR: 7–10 days). Extraordinary short interval of 1 day from oxygen therapy to respiratory support indicates a rapid worsening of hypoxemia in COVID-19. The duration of respiratory support was 11 days (IQR: 5–18 days), and the overall mortality rate in ICU was 25.0%. Days of respiratory support were 19 vs 7 days and ICU mortality was 71.4% vs 0.0% in the failed to discontinue and successfully discontinued groups, respectively (significant difference, both *p*-value < 0.001).

**Table 1.**
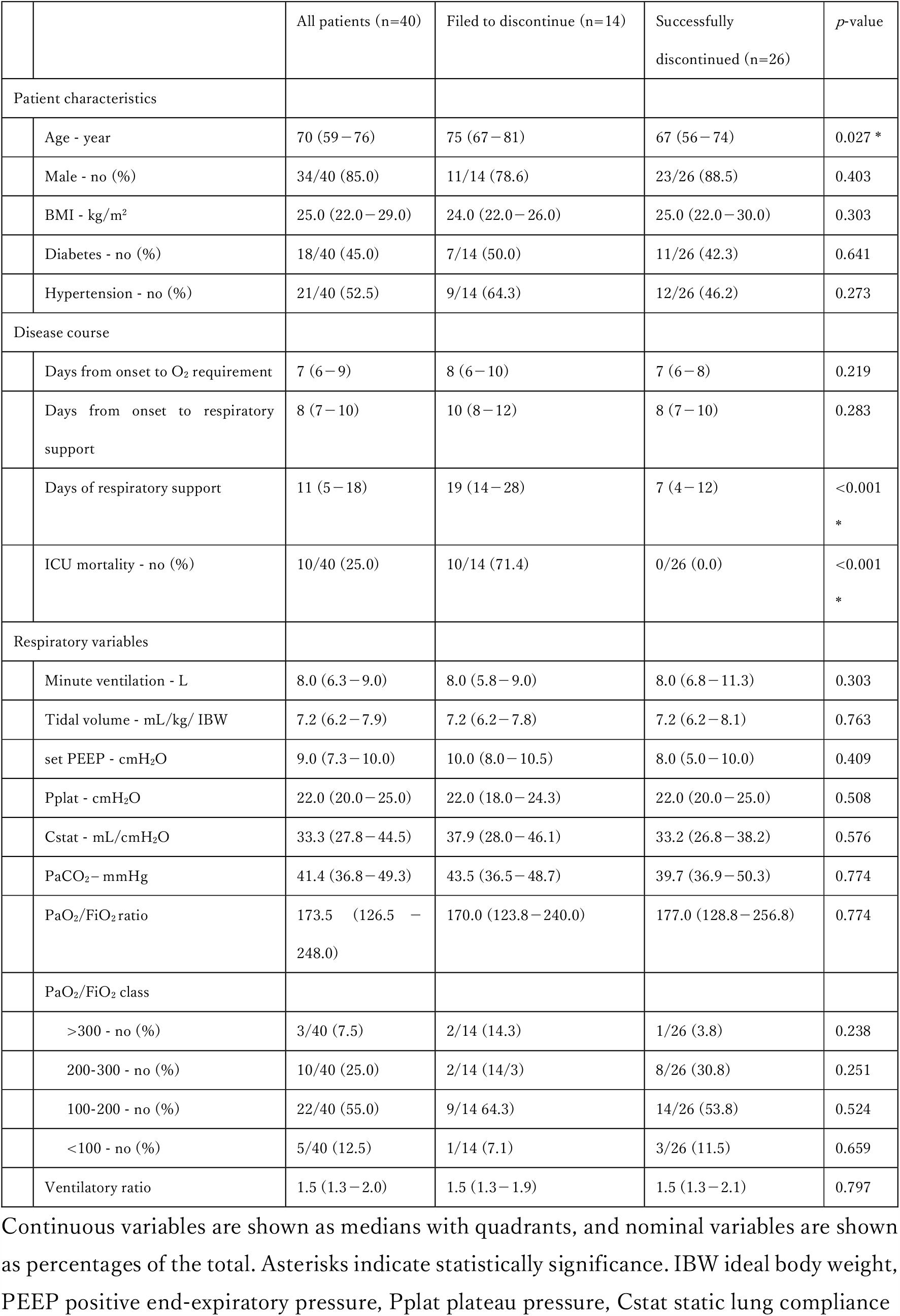
Demographic data, disease course, and respiratory variables.

|  |  | All patients (n=40) | Filed to discontinue (n=14) | Successfully discontinued (n=26) | p-value |
| --- | --- | --- | --- | --- | --- |
| Patient characteristics |  |  |  |  |  |
|  | Age - year | 70 (59–76) | 75 (67–81) | 67 (56–74) | 0.027 * |
|  | Male - no (%) | 34/40 (85.0) | 11/14 (78.6) | 23/26 (88.5) | 0.403 |
|  | BMI - kg/m<sup>2</sup> | 25.0 (22.0–29.0) | 24.0 (22.0–26.0) | 25.0 (22.0–30.0) | 0.303 |
|  | Diabetes - no (%) | 18/40 (45.0) | 7/14 (50.0) | 11/26 (42.3) | 0.641 |
|  | Hypertension - no (%) | 21/40 (52.5) | 9/14 (64.3) | 12/26 (46.2) | 0.273 |
| Disease course |  |  |  |  |  |
|  | Days from onset to O<sub>2</sub> requirement | 7 (6–9) | 8 (6–10) | 7 (6–8) | 0.219 |
|  | Days from onset to respiratory support | 8 (7–10) | 10 (8–12) | 8 (7–10) | 0.283 |
|  | Days of respiratory support | 11 (5–18) | 19 (14–28) | 7 (4–12) | <0.001<br>* |
|  | ICU mortality - no (%) | 10/40 (25.0) | 10/14 (71.4) | 0/26 (0.0) | <0.001<br>* |
| Respiratory variables |  |  |  |  |  |
|  | Minute ventilation - L | 8.0 (6.3–9.0) | 8.0 (5.8–9.0) | 8.0 (6.8–11.3) | 0.303 |
|  | Tidal volume - mL/kg/ IBW | 7.2 (6.2–7.9) | 7.2 (6.2–7.8) | 7.2 (6.2–8.1) | 0.763 |
|  | set PEEP - cmH<sub>2</sub>O | 9.0 (7.3–10.0) | 10.0 (8.0–10.5) | 8.0 (5.0–10.0) | 0.409 |
|  | Pplat - cmH<sub>2</sub>O | 22.0 (20.0–25.0) | 22.0 (18.0–24.3) | 22.0 (20.0–25.0) | 0.508 |
|  | Cstat - mL/cmH<sub>2</sub>O | 33.3 (27.8–44.5) | 37.9 (28.0–46.1) | 33.2 (26.8–38.2) | 0.576 |
|  | PaCO<sub>2</sub> - mmHg | 41.4 (36.8–49.3) | 43.5 (36.5–48.7) | 39.7 (36.9–50.3) | 0.774 |
|  | PaO<sub>2</sub>/FiO<sub>2</sub> ratio | 173.5 (126.5 – 248.0) | 170.0 (123.8–240.0) | 177.0 (128.8–256.8) | 0.774 |
|  | PaO<sub>2</sub>/FiO<sub>2</sub> class |  |  |  |  |
|  | >300 - no (%) | 3/40 (7.5) | 2/14 (14.3) | 1/26 (3.8) | 0.238 |
|  | 200-300 - no (%) | 10/40 (25.0) | 2/14 (14/3) | 8/26 (30.8) | 0.251 |
|  | 100-200 - no (%) | 22/40 (55.0) | 9/14 64.3) | 14/26 (53.8) | 0.524 |
|  | <100 - no (%) | 5/40 (12.5) | 1/14 (7.1) | 3/26 (11.5) | 0.659 |
|  | Ventilatory ratio | 1.5 (1.3–2.0) | 1.5 (1.3–1.9) | 1.5 (1.3–2.1) | 0.797 |

Continuous variables are shown as medians with quadrants, and nominal variables are shown as percentages of the total. Asterisks indicate statistical significance. IBW ideal body weight, PEEP positive end-expiratory pressure, Pplat plateau pressure, Cstat static lung compliance

Age was significantly greater in the failed to discontinue group than the successfully discontinued group (median, 75 vs 67 years respectively, *p*-value = 0.027). There was a predominance of males (34/40, 85.0%). Median BMI was 25 kg/m^2^ (IQR, 22–29 kg/m^2^). The incidence of the comorbidities of diabetes (18/40, 45.0%) and hypertension (21/40, 52.5%) in all patients in the study were higher than the prevalence in Japanese men (13.8% and 23.0%, respectively adjusted by age)(12).

The median values of respiratory parameters at 2 hours (day 0) after initiation of respiratory support in all patients were as follows: MV, 8.0 L (IQR, 6.3–9.0 L); TV, 7.2 mL/kg (IQR, 6.2–7.9 mL/kg); set PEEP, 9.0 cmH_2_O (IQR, 7.3–10.0 cmH_2_O); and Pplat, 22.0 cmH_2_O (IQR, 20.0–25.0 cmH_2_O). Lung static compliance was 33.3 mL/mmH_2_O (IQR, 27.8–44.5 mL/mmH_2_O), which is slightly lower than the 40 mL/cmH_2_O in the Berlin definition (13). PaCO_2_ was 41.4 mmHg (IQR, 36.8–49.3 mmHg), and there was no apparent hypercapnia. P/F ratio was 173.5 (IQR, 126.5–248.0), and ARDS was most frequently moderate in severity. VR was 1.5 (IQR, 1.3–2.0), higher than the value of 1. The normal value of VR is around 1, but increases with worsening ventilation mechanics, excess CO_2_ production, or both (7). There was no significant difference in any of these respiratory parameters between the two groups.

Figure 1 shows the longitudinal VR values from 2 hours (day 0) of respiratory support to day 6. VR was stable until around day 2 after initiation of respiratory support in both groups. In the failed to discontinue group, VR increased on day 3. VR was significantly higher in the failed to discontinue group than in the successfully discontinued group on day 4 after respiratory support (*p*-value = 0.027) (Figure 1a). Throughout the observation period there was no apparent trend in the change in PaCO_2_ or MV, which are both used to calculate VR (Figure 1b, c).

**Figure 1.**
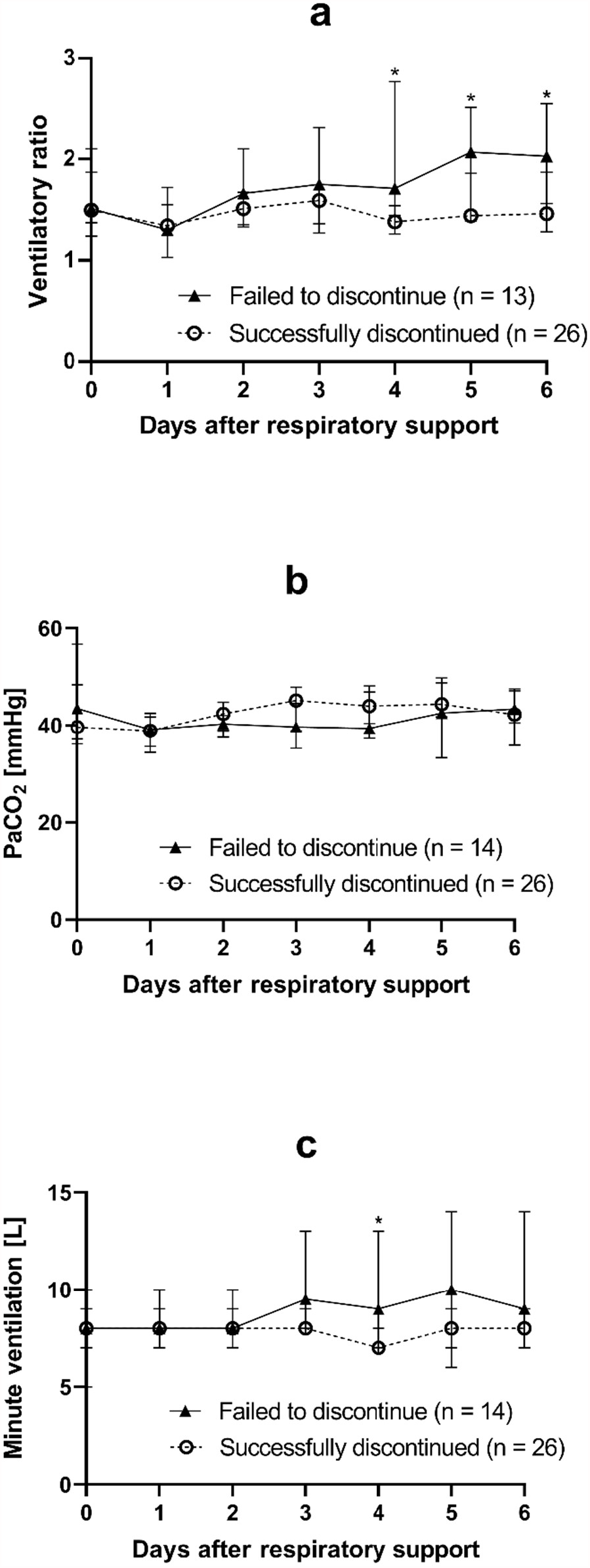
Dynamics of ventilatory ratio, PaCO_2_, and minute ventilation from 2 hours (day 0) to day 6 after initiation of respiratory support. Longitudinal daily values of ventilatory ratio (VR) (a), PaCO_2_ (b), and minute ventilation (c) from 2 hours (day 0) after initiation of respiratory support to the following 6 days. The failed to discontinue group presented as the solid triangles and solid lines, and the successfully discontinued group presented as the opened circles and the dashed lines. Each marker placed on the median value, and each error bar meant the 95% confidential interval. Statistical analysis determined the differences between the failed to discontinue and successfully discontinued groups on each day by Mann-Whitney test. ^*^ *p*-value < 0.05

Figure 2 shows ROC curves for VR on days 0-4 after initiation of respiratory support. Differences in VR values are seen between the two groups with the progression of time. The area under the ROC curve (AUC) maximized on day 4 was 0.748, which indicates moderate accuracy (statistically significant, *p*-value = 0.019) (Table 2). The cut-off value for maximum diagnostic performance with a sensitivity of 0.667 and a specificity of 0.762 was VR > 1.56. Using VR > 1.56 as the threshold in the present cohort, the failed to discontinue (positive) predictive value was 0.667 (true positive 8 vs false positive 4) and the successfully discontinued (negative) predictive value was 0.810 (true negative 17 vs false negative 4) on day 4.

**Table 2.** The parameters of the area under ROC curve on each day after initiation of respiratory support.

| After initiation of respiratory support | Area | Standard error | 95% CI | *p*-value |
| --- | --- | --- | --- | --- |
| 2 hours (day 0) | 0.527 | 0.096 | 0.339 to 0.714 | 0.789 |
| Day 1 | 0.520 | 0.112 | 0.301 to 0.738 | 0.854 |
| Day 2 | 0.659 | 0.100 | 0.463 to 0.854 | 0.120 |
| Day 3 | 0.678 | 0.100 | 0.483 to 0.872 | 0.074 |
| Day 4 | 0.748 | 0.091 | 0.570 to 0.927 | 0.019 * |

**Figure 2.**
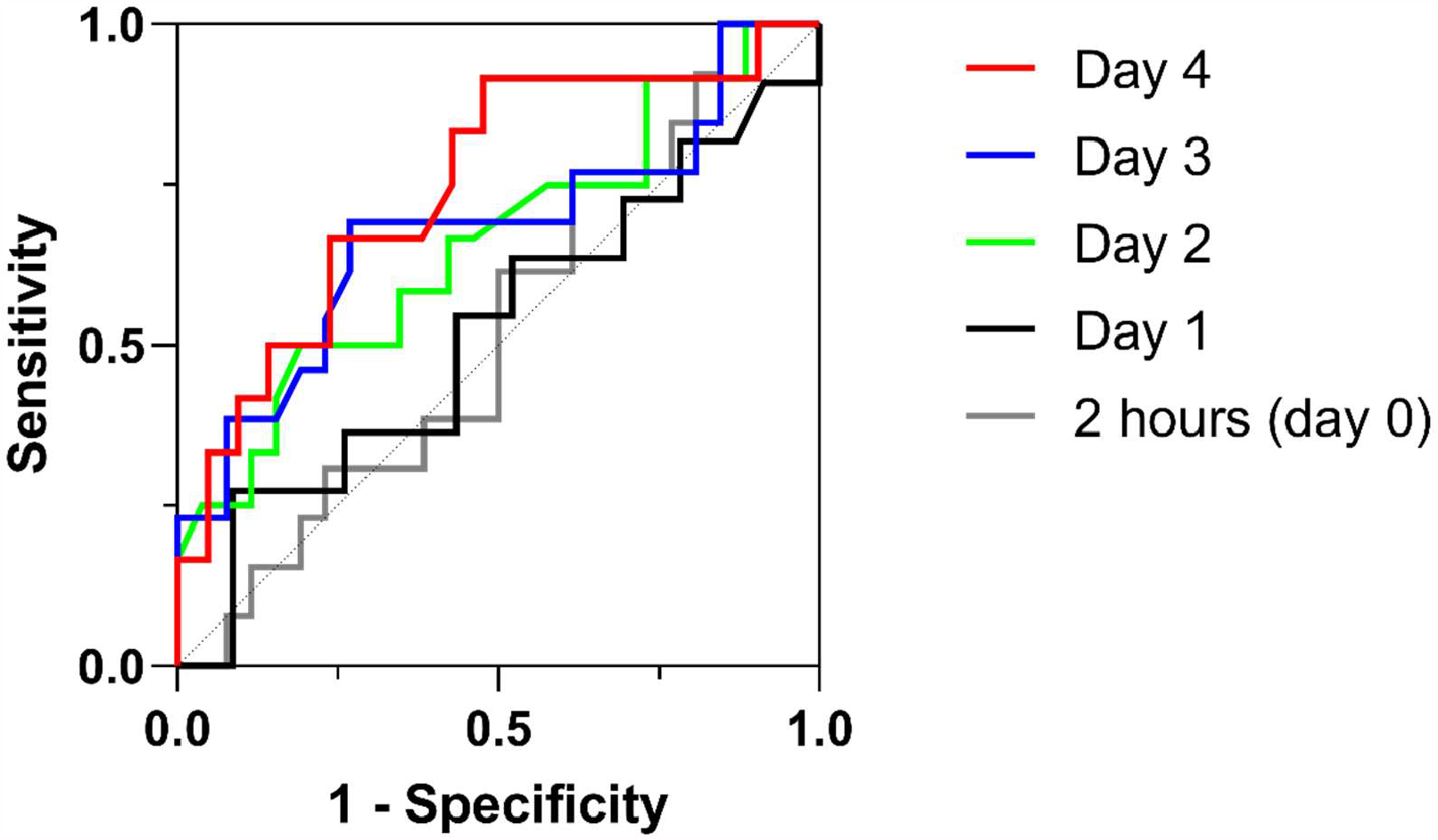
Receiver operating characteristics (ROC) curves for VR on days 0–4 after initiation of respiratory support. The prognostic ability of VR on the individual day was evaluated by ROC curve analysis. A failed to discontinue outcome was analyzed as positive. The area under the ROC curve (AUC) maximized on day 4 after initiation of respiratory support at 0.748, which indicates moderate accuracy. The standard error was 0.091, 95% confidential interval was 0.570 to 0.927, and the *p*-value was 0.019 (Table 2). The cut-off value for a maximum diagnostic performance with a sensitivity of 0.667 and a specificity of 0.762 was VR > 1.56.

## Discussion

This study monitored longitudinal VR in patients with severe COVID-19 who required respiratory support in our ICU with aim of determining whether VR could stratify the severity of COVID-19 and predicting successfully discontinued prognosis. The main findings were that VR on day 4 after initiation of respiratory support was higher than in the failed to discontinue group than the successfully discontinued group, and that a threshold of VR < 1.56 was predictive of successful discontinuation with a predictive value of 0.810.

Pulmonary pathology of COVID-19 can be subdivided into four main morphological stages: [1] early stage (day 0–1) with edema, incipient epithelial damage, and capillaritis, [2] exudative DAD stage (days 1–7), [3] organizing stage (1 to several weeks), and [4] fibrotic DAD stage (weeks to months) (14). Pre-symptomatic histology observed in surgical sections obtained from patients who were later found to have COVID-19 included proteinaceous exudate, focal reactive hyperplasia of pneumocytes, alveolar hemorrhage, and inflammatory cellular infiltration (15,16). Autopsy of acute COVID-19 patients within 20 days from symptom onset revealed exudative DAD, massive capillary congestion, and pulmonary embolism (17). The time point of the increase in VR found in the present study coincides with the stage of exudative DAD. Therefore, the increased VR on day 4 after initiation of respiratory support may indicate the severity of DAD.

From the immunological perspective, neutralizing antibody and T-cell responses play essential roles in eliminating SARS-CoV-2 from the body (18,19). In previous studies, patients who experience mild symptoms eliminated the virus within 15 days, whereas patients with moderate and severe symptoms take much longer to eliminate the virus from the upper respiratory tract (18). Peak neutralizing activity of virus-specific antibodies was achieved within 9 – 15 days after symptom onset, and patients with moderate/severe symptoms exhibited more robust responses than those with mild disease. Higher frequencies of IFN-γ-secreting T cells at around day 15 from symptom onset were present in mild, but not in moderate/severe COVID-19 patients (18). Considering interval from symptom onset to respiratory support averaged to 8 days, the time point of the VR increase on day 4 after respiratory support in the failed to discontinue group coincided with the immunological turning point at around days 10–15 from symptom onset. The increased VR on day 4 after respiratory support might reflect the immunological dynamics in patients with severe COVID-19.

The ROC curve of VR on day 4 after initiation of respiratory support was predictive of successful discontinuation from respiratory support with a high predictive value in this cohort. Therefore, VR values may indicate the effectiveness of current treatment. Moreover, the stratification of severe COVID-19 according to VR may enable consideration of the appropriate therapy on an individual basis, such as the decision whether to continue with aggressive COVID-19-specific therapies. VR has a potential to be a candidate predictor for improving predictive abilities of current risk prediction models for ICU patients with COVID-19 (20).

In contrast to the present study, some previous reports (21–23) that have used VR to assess the clinical condition of patients with COVID-19 have concluded that VR is a minimally useful indicator. The reason for the discrepancy in results may be that these previous studies calculated VR only once, on the day that respiratory support was initiated. As demonstrated in the present study, a significant difference in VR values between the failed to discontinue and successfully discontinued groups was apparent on day 4 after initiation of respiratory support, and the difference was a predictor of successfully discontinued prognosis. VR being an easily calculated bedside friendly parameter, frequent assessment and longitudinal monitoring is feasible and potentially useful in understanding the severity status of COVID-19 patients.

This study has some limitations. The most appropriate VR value to be used as the threshold requires further evaluation. This study only used data up to day 6 of respiratory support. Data were not available after day 7 because a large number of patients were withdrawn from respiratory support at the time. Finally, we cannot rule out the possibility of selection bias because we included only a small number of patients collected at a single institution.

## Conclusions

Increased VR values on day 4 after initiation of respiratory support is predictive of successful discontinuation in patients with severe COVID-19. Day 4 after respiratory support coincides with the time point of the exudative DAD stage in pulmonary pathology, and the turning point of both neutralizing activity and T-cell responses in immunological dynamics. Longitudinal VR values after respiratory support may be a suitable practical index for stratifying patients with severe COVID-19 and predict successfully discontinued outcome.

## Data Availability

The datasets used and analyzed during the current study are available in Microsoft Excel files titled 'Supplement Table 1－4'.

## List of abbreviations

ACE2: angiotensin-converting enzyme 2
ARDS: acute respiratory distress syndrome
AUC: area under ROC curve
BMI: body mass index
COVID-19: Coronavirus disease 2019
DAD: diffuse alveolar damage
Cstat: static lung compliance
FiO2: the fraction of inspired oxygen
IBW: ideal body weight
ICU: intensive care unit
MERS-CoV: middle east respiratory syndrome coronavirus
MV: minute ventilation
PaCO_2_: the partial pressure of carbon dioxide
PaO_2_: the partial pressure of oxygen
PCR: polymerase chain reaction
PEEP: positive end-expiratory pressure
Pplat: plateau pressure
P/F: PaO_2_ / FiO_2_
ROC: receiver operating characteristic
SARS-CoV: severe acute respiratory syndrome coronavirus
SARS-CoV-2: severe acute respiratory syndrome coronavirus 2
V_D_/V_T_: dead space fraction
VR: ventilatory ratio
VT: tidal volume

## Declarations

### Ethics approval and consent to participate

This study was conducted under the approval of the Institutional Review Board of Osaka City University (#2020-003) and the Clinical Research Ethics Committee of the Osaka Municipal Hospital Organization (#2005020), Osaka, Japan.

### Consent for publication

Not applicable.

### Availability of data and materials

The datasets used and analyzed during the current study are available in Microsoft Excel files titled ‘Supplement Table 1–4’.

### Competing interests

The authors declare that they have no competing interests.

### Funding

This work was funded by Japan Agency for Medical Research and Development (AMED) under Grant number JP20wm0125003 (Yasutoshi Kido), JP20he1122001 (Yasutoshi Kido), JP20nk0101627 (Yasutoshi Kido), and JP20jk0110021 (Yu Nakagama). This work was also supported by JSPS KAKENHI Grans Number JP21441824 (Natsuko Kaku). We also receive the COVID-19 Private Fund (to the Shinya Yamanaka laboratory, CiRA, Kyoto University). We received support from Osaka City University’s “Special Reserves” fund for COVID-19.

### Authors’ contributions

N Kaku, M Shirano, and Y Kido designed the concept of the study. N Kaku, S Shinomiya, K Yamazaki, K Shimazu, and Y Maehata, and R Morita performed the data acquisition. N Kaku, Y Nakagama, Y Nitahara, and Y Kido assessed the quality of the study and performed the analysis and interpretation. N Kaku, Y Nakagama, and Y Kido wrote the manuscript, and the other authors made substantial revisions and edits. All authors read and approved the final manuscript.

## Acknowledgements

I am grateful to FORTE science communications for the manuscript revision. I would like to thank the members of Osaka City General Hospital, where the main results of this paper were obtained, especially Dr. T Nishida and Dr. T Shigemoto. I would also like to take this opportunity to thank Dr. T Akamine, Dr. E Kataoka-Nakamatsu, Dr. R Uchida, and Dr. Y Morimoto for their kind supports.

---

Supplement Table 1. Detailed information of demographic data, disease course, and respiratory variables.

Supplement Table 2. Raw data of ventilatory ratio from 2 hours (day 0) to day 6 after initiation of respiratory support.

Supplement Table 3. Raw data of PaCO_2_ from 2 hours (day 0) to day 6 after initiation of respiratory support.

Supplement Table 4. Raw data of minute ventilation from 2 hours (day 0) to day 6 after initiation of respiratory support.

## Notes

### Competing Interest Statement

The authors have declared no competing interest.

